# Exercise-Derived Hemodynamics and 6-Minute Walk Distance in Hypertension

**DOI:** 10.64898/2026.07.30.26359378

**Authors:** Ting Yu, Min Wang, Meng Liu, Ye Zhou, Ying Cao, Minzhe Shi, Yuntao Li, Mei Hong, Guozhong Ji

## Abstract

**Background:** Patients with hypertension commonly exhibit reduced exercise capacity. Impedance cardiography (ICG) enables continuous, non-invasive assessment of exercise-derived hemodynamic parameters; however, the relative associations of different ICG-derived parameters with exercise capacity remain unclear.

**Methods:** In this retrospective cross-sectional study, 211 patients with hypertension who completed both ICG assessment and the six-minute walk test (6MWT) were enrolled. Exercise-derived hemodynamic parameters, including maximum heart rate (HRmax), stroke volume (SV), cardiac output (CO), and systemic vascular resistance (SVR), were continuously measured using the PhysioFlow® system. Univariable and multivariable linear regression analyses were performed to evaluate the associations between ICG-derived parameters and six-minute walk distance (6MWD). HRmax tertile and subgroup analyses were subsequently conducted.

**Results:** In the final multivariable model, age (β = −2.76, 95% CI, −3.85 to −1.67; *P* < 0.001), male sex (β = 33.50, 95% CI, 8.40–58.60; *P* = 0.009), and HRmax (β = 1.26, 95% CI, 0.69–1.83; *P* < 0.001) were independently associated with 6MWD. 6MWD increased progressively across HRmax tertiles (*P* for trend < 0.001). The positive association between HRmax and 6MWD remained consistent across all prespecified and exploratory subgroups.

**Conclusions:** Among multiple exercise-derived ICG hemodynamic parameters, HRmax demonstrated the strongest and most consistent independent association with exercise capacity in patients with hypertension. These findings suggest that HRmax may represent the most informative dynamic hemodynamic parameter during exercise and that combining ICG with the 6MWT may provide a simple and accessible complementary approach for evaluating exercise capacity.

## Introduction

Hypertension is one of the most common chronic cardiovascular diseases worldwide and represents a major risk factor for heart failure, cardiovascular events, and all-cause mortality^[1]^. Increasing evidence suggests that patients with hypertension exhibit reduced exercise capacity even in the absence of overt heart failure. Impaired exercise capacity not only adversely affects quality of life but also reflects impaired circulatory reserve^[2–4]^. Therefore, early identification of reduced exercise capacity and its associated factors is of considerable importance for risk stratification and individualized management in patients with hypertension.

Various objective techniques have been used to characterize impaired exercise capacity in patients with hypertension. Cardiopulmonary exercise testing (CPET) studies have demonstrated reduced peak oxygen uptake (peak VO₂), accompanied by abnormal hemodynamic and metabolic responses, in patients with hypertension^[2]^.Stress echocardiographic studies have shown that diastolic dysfunction, impaired left atrial reserve, and altered myocardial function are associated with reduced exercise capacity^[3,4]^. In addition, studies using impedance cardiography (ICG) have demonstrated that exercise-derived ICG hemodynamic parameters are closely associated with exercise capacity^[5]^. Although impaired exercise capacity in patients with hypertension has been demonstrated using multiple assessment techniques, the relative contributions of different hemodynamic parameters to exercise capacity remain incompletely understood.

CPET is considered the gold standard for the assessment of exercise capacity because it provides a comprehensive evaluation of cardiopulmonary function and metabolic reserve. However, its widespread use in primary healthcare settings and routine clinical practice is limited by the high cost of equipment, procedural complexity, and the need for specialized expertise^[6]^. The six-minute walk test (6MWT) is a simple, safe, and reproducible submaximal exercise test that provides a practical assessment of functional exercise capacity. The American Thoracic Society (ATS) recommends the 6MWT as a standardized method for evaluating functional exercise capacity^[7]^. Furthermore, previous studies have shown that exercise-derived ICG hemodynamic parameters obtained during the 6MWT can predict peak oxygen uptake (peak VO₂) measured by CPET, suggesting that this approach may provide a reasonable surrogate measure of aerobic exercise capacity^[8]^.

ICG is a non-invasive and convenient technique for hemodynamic monitoring that enables real-time assessment of heart rate (HR), stroke volume (SV), cardiac output (CO), and systemic vascular resistance (SVR). Previous studies have suggested that exercise-derived ICG hemodynamic parameters better reflect circulatory reserve than measurements obtained at rest^[5,9]^. However, it remains unclear whether different exercise-derived hemodynamic parameters have comparable associations with exercise capacity. Therefore, the present study combined ICG with the 6MWT and used six-minute walk distance (6MWD) as the indicator of exercise capacity to systematically evaluate the associations between multiple exercise-derived ICG hemodynamic parameters and 6MWD in patients with hypertension, with the aim of identifying the parameter most closely associated with exercise capacity.

## Methods

### Study Population

This retrospective cross-sectional study consecutively enrolled 211 patients with hypertension who underwent non-invasive cardiac function assessment and 6MWT at The Second Affiliated Hospital of Nanjing Medical University between January 2025 and January 2026.

The diagnosis of hypertension was established according to the 2024 European Society of Cardiology (ESC) Guidelines for the Management of Elevated Blood Pressure and Hypertension^[1]^. The inclusion criteria were as follows: (1) age ≥18 years; (2) a confirmed diagnosis of hypertension; (3) completion of ICG and the 6MWT; and (4) complete clinical data.

The exclusion criteria were as follows: (1) acute coronary syndrome or acute decompensated heart failure; (2) severe cardiac arrhythmias; (3) severe valvular heart disease; (4) severe hepatic or renal dysfunction; (5) malignant tumors or active infection; (6) inability to complete the 6MWT because of musculoskeletal disorders, neurological disorders, or other conditions; and (7) incomplete clinical data.

### Ethics Statement

The study was approved by the Ethics Committee of The Second Affiliated Hospital of Nanjing Medical University (Approval No. 2026-KY-285-01). The study was conducted in accordance with the principles of the Declaration of Helsinki.

### Clinical Data Collection and Laboratory Measurements

Demographic characteristics, comorbidities, laboratory measurements, and echocardiographic parameters were collected for all participants. Demographic data included age and sex. Comorbidities included coronary heart disease (CHD) and congestive heart failure (CHF).

Laboratory measurements included C-reactive protein (CRP), neutrophil-to-lymphocyte ratio (NLR), and estimated glomerular filtration rate (eGFR). The NLR was defined as the ratio of the absolute neutrophil count to the absolute lymphocyte count in peripheral blood. The eGFR was calculated using the Chronic Kidney Disease Epidemiology Collaboration (CKD-EPI) equation^[10]^.

Left ventricular ejection fraction (LVEF) was assessed by transthoracic echocardiography.

### Impedance Cardiography and Six-Minute Walk Test

Non-invasive hemodynamic monitoring was performed using the PhysioFlow® impedance cardiography system ^[11]^. Hemodynamic parameters were continuously recorded at rest and during exercise, including heart rate (HR), maximum heart rate (HRmax), stroke volume (SV), maximum stroke volume (SVmax), cardiac output (CO), and systemic vascular resistance (SVR).

The 6MWT was performed according to ATS guidelines. Participants were instructed to walk as far as possible for 6 minutes along a flat corridor. 6MWD (m) was recorded as the primary measure of exercise capacity^[7]^.

### Statistical Analysis

The normality of continuous variables was assessed using the Shapiro–Wilk test. Normally distributed variables were presented as mean ± standard deviation, non-normally distributed variables as median (interquartile range), and categorical variables as number (percentage). Missing values were imputed using the median. Univariable linear regression analysis was first performed to identify factors associated with 6MWD. Variables with *P* < 0.10 in the univariable analysis, together with clinically relevant variables, were entered into multivariable linear regression models. Multiple candidate models were constructed, and the stability of key regression coefficients was evaluated. Model fit was assessed using the adjusted coefficient of determination (adjusted R²), Akaike information criterion (AIC), and Bayesian information criterion (BIC). Multicollinearity was evaluated using the variance inflation factor (VIF). Model robustness was assessed using 10-fold cross-validation, and the final model was selected on the basis of statistical performance and clinical interpretability.

To evaluate the linear association between HRmax and 6MWD, a quadratic term for HRmax was incorporated into the final multivariable model. Scatter plots were generated to visualize the relationship between HRmax and 6MWD.

Participants were further categorized into HRmax tertiles according to the distribution of HRmax. Multivariable linear regression analysis was performed to examine the association between HRmax tertiles and 6MWD. A test for trend (*P* for trend) was conducted by modeling HRmax tertiles as an ordinal variable. In addition, boxplots were generated to illustrate the distribution of 6MWD across HRmax tertiles. Prespecified subgroup analyses were performed according to sex, age, and CHD, whereas exploratory subgroup analyses were conducted according to NLR, eGFR, and body mass index (BMI). Effect modification was evaluated by including interaction terms in the regression models, and the results of subgroup analyses were presented as forest plots.

All statistical analyses were performed using R software (version 4.6.0). All tests were two-sided, and a *P* value < 0.05 was considered statistically significant.

## Results

### Baseline Characteristics

A total of 211 patients with hypertension were included in this study. All participants successfully completed the 6MWT, and no adverse events leading to test termination occurred during the procedure.

The mean age of the study population was 62.0 years, and 108 participants (51.18%) were male. The median HRmax was 114.0 bpm (interquartile range [IQR], 103.0–126.0 bpm), and the median 6MWD was 540 m (IQR, 480–590 m). The prevalence of CHD and CHF was 35.55% and 2.37%, respectively. The remaining baseline characteristics, hemodynamic parameters, and laboratory measurements are presented in Table 1.

**Table 1.**
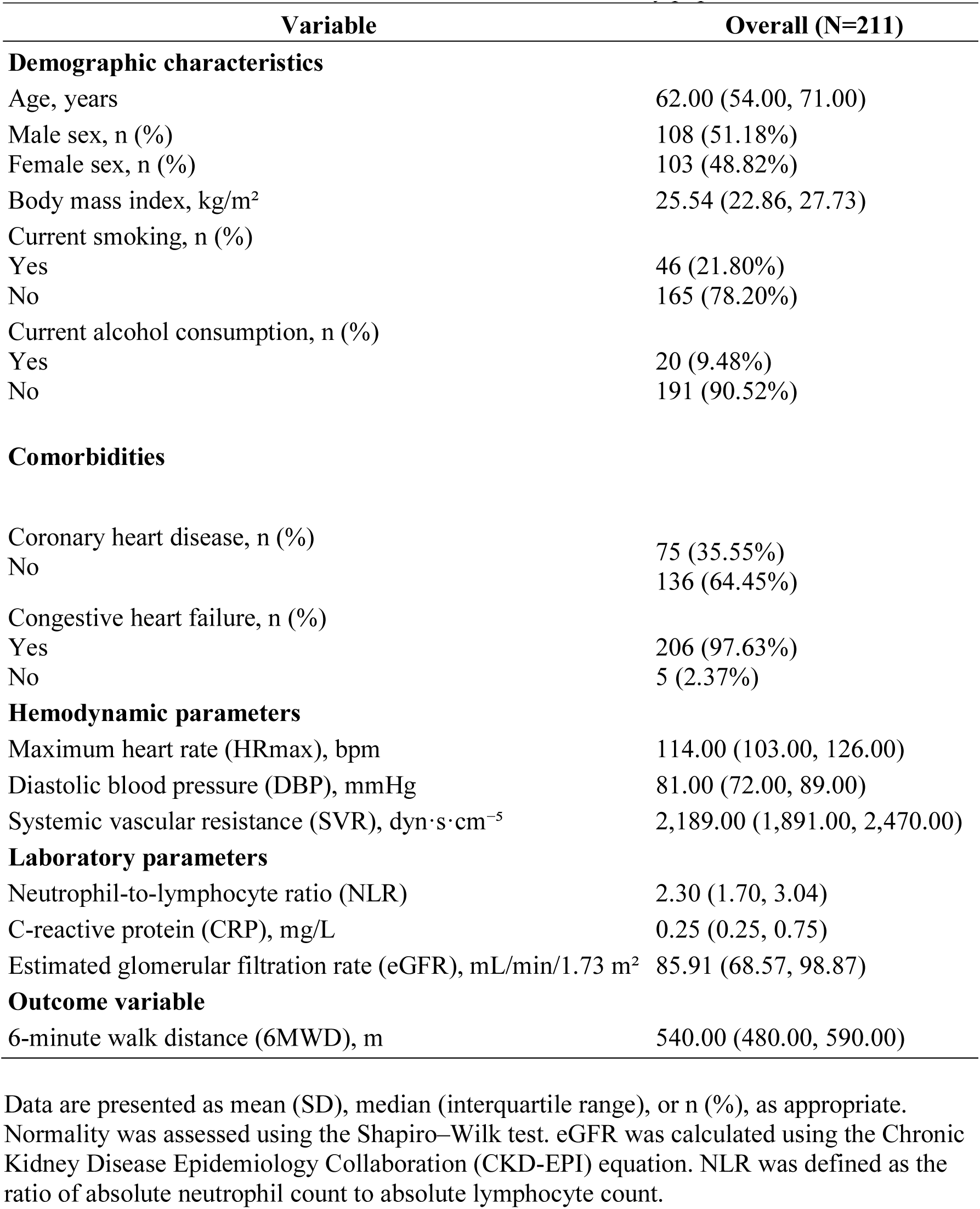
Baseline characteristics of the study population.

### Association Between Hemodynamic Parameters and 6MWD

In the univariable analysis, sex, CHD, CHF, age, HRmax, DBP, SVR, NLR, CRP, and eGFR were significantly associated with 6MWD. The results of model comparison, multicollinearity assessment, and cross-validation are presented in Supplementary Tables S1–S3.

In the final multivariable model, age, sex, and HRmax remained independently associated with 6MWD. Specifically, each 1-year increase in age was associated with a 2.76-m decrease in 6MWD (β = −2.76; 95% CI, −3.85 to −1.67; *P* < 0.001), whereas male participants walked, on average, 33.50 m farther than female participants (β = 33.50; 95% CI, 8.40 to 58.60; *P* = 0.009). HRmax was positively associated with 6MWD, with each 1-bpm increase in HRmax being associated with a 1.26-m increase in 6MWD (β = 1.26; 95% CI, 0.69 to 1.83; *P* < 0.001) (Table 2).

**Table 2.** Univariable and multivariable analyses.

| Variable | Univariate $\beta$ (95%CI) | *P* | Multivariable $\beta$ (95%CI) | *P* |
| --- | --- | --- | --- | --- |
| Sex | 30.78(7.50,54.06) | 0.010 | 33.50 (8.41, 58.59) | 0.009 |
| CHD | - 48.54 ( - 72.34, - 24.75) | <0.001 | - 18.83 ( - 39.34,1.69) | 0.072 |
| CHF | - 137.93 (213.35, - 62.51) | <0.001 | - 61.75 ( - 125.63,2.14) | 0.058 |
| Age | - 3.77 ( - 4.59, - 2.96) | <0.001 | - 2.76 ( - 3.85, - 1.67) | <0.001 |
| HRmax | 2.19 (1.60, 2.77) | <0.001 | 1.26 (0.69,1.83) | <0.001 |
| DBP | 1.51(0.65,2.52) | 0.001 | - 0.85( - 1.79,0.08) | 0.074 |
| SVR | 0.022(<0.01,0.04) | 0.041 | 0.016(<-0.01,0.035) | 0.089 |
| NLR | - 16.329( - 26.50, - 6.16) | 0.002 | - 6.474( - 15.36,2.41) | 0.152 |
| CRP | - 1.553( - 3.21,0.11) | 0.067 | - 0.661( - 2.02,0.70) | 0.338 |
| eGFR | 0.920(0.34,1.50) | 0.002 | 0.082( - 0.64,0.81) | 0.824 |

$\beta$ coefficients and 95% confidence intervals (95% CI) were estimated using linear regression analysis. Variables with $P < 0.10$ in univariable analyses and clinically relevant variables were included in the multivariable model.
Abbreviations:CHD, coronary heart disease;CHF, congestive heart failure;HRmax, maximum heart rate;DBP, diastolic blood pressure;SVR, systemic vascular resistance;NLR, neutrophil-to-lymphocyte ratio;CRP, C-reactive protein;eGFR, estimated glomerular filtration rate.

### Linearity Analysis of the Association Between HRmax and 6MWD

To assess the linearity of the association between HRmax and 6MWD, a quadratic term for HRmax was added to the final multivariable model. The quadratic term of HRmax was not statistically significant (*P* = 0.629), indicating no evidence of a nonlinear association (Supplementary Table S4). The scatter plot demonstrated a positive linear association between HRmax and 6MWD (Figure 1).

**Figure 1.**
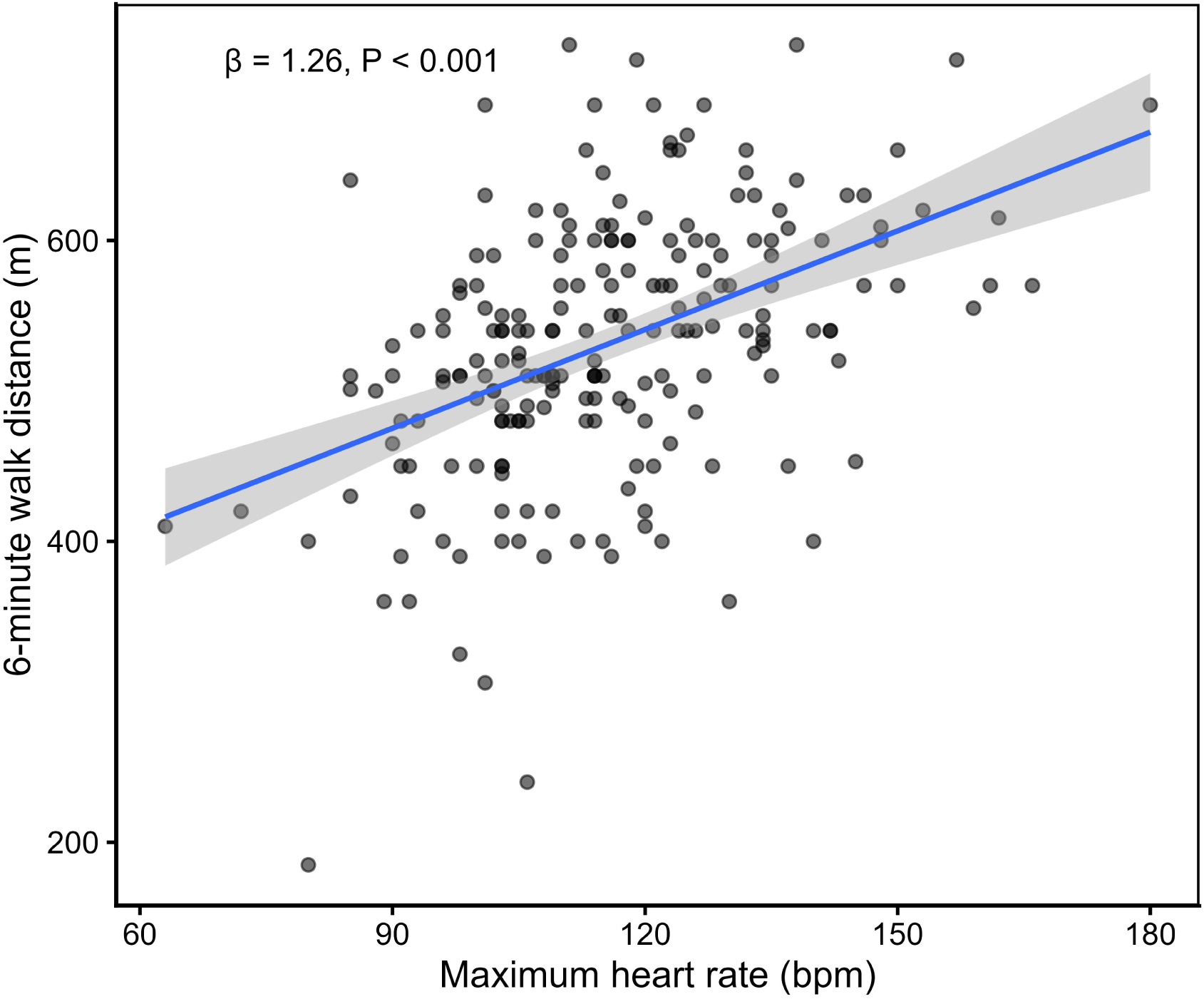
Association between maximum heart rate and 6-minute walk distance. A positive linear association was observed between HRmax and 6MWD. The solid line represents the fitted linear regression line and the shaded area represents the 95% confidence interval. Abbreviations:HRmax, maximum heart rate;6MWD, six-minute walk distance.

### Association Between HRmax Tertiles and 6MWD

Participants were categorized into three groups according to HRmax tertiles: T1 (lowest tertile), T2 (middle tertile), and T3 (highest tertile). After adjustment for potential confounders, 6MWD was significantly greater in both the T2 and T3 groups than in the T1 group.Compared with the T1 group, the T2 group had a 27.06-m higher 6MWD (β = 27.06; 95% CI, 3.43–50.69; *P* = 0.025), whereas the T3 group had a 50.23-m higher 6MWD (β = 50.23; 95% CI, 24.81–75.64; *P* < 0.001). The test for trend demonstrated a progressive increase in 6MWD across HRmax tertiles (*P* for trend < 0.001) (Table 3).

**Table 3.** Maximum heart rate tertile regression.

| **HRmax tertiles** | **$\beta$ (95% CI)** | ***P* value** |
| --- | --- | --- |
| T1 (Reference) | Ref | — |
| T2 | 27.06(3.43, 50.69) | 0.025 |
| T3 | 50.23(24.81, 75.64) | <0.001 |
| P for trend | 25.12(12.45, 37.80) | <0.001 |

T1 served as the reference group. Adjusted for sex, CHD, CHF, age, DBP, SVR, NLR, CRP, and eGFR.
Abbreviations: CHD, coronary heart disease; CHF, congestive heart failure; DBP, diastolic blood pressure; SVR, systemic vascular resistance; NLR, neutrophil-to-lymphocyte ratio; CRP, C-reactive protein; eGFR, estimated glomerular filtration rate.

Baseline characteristics stratified by HRmax tertiles are presented in Supplementary Table S5, and the distribution of 6MWD across HRmax tertiles is shown in Supplementary Figure S1.

### Subgroup and Sensitivity Analyses

The positive association between HRmax and 6MWD remained consistent across all prespecified and exploratory subgroups (Figure 2). No significant effect modification was observed according to sex, age group, CHD, NLR, eGFR, or BMI (all *P* for interaction > 0.05) (Supplementary Table S6).

**Figure 2.**
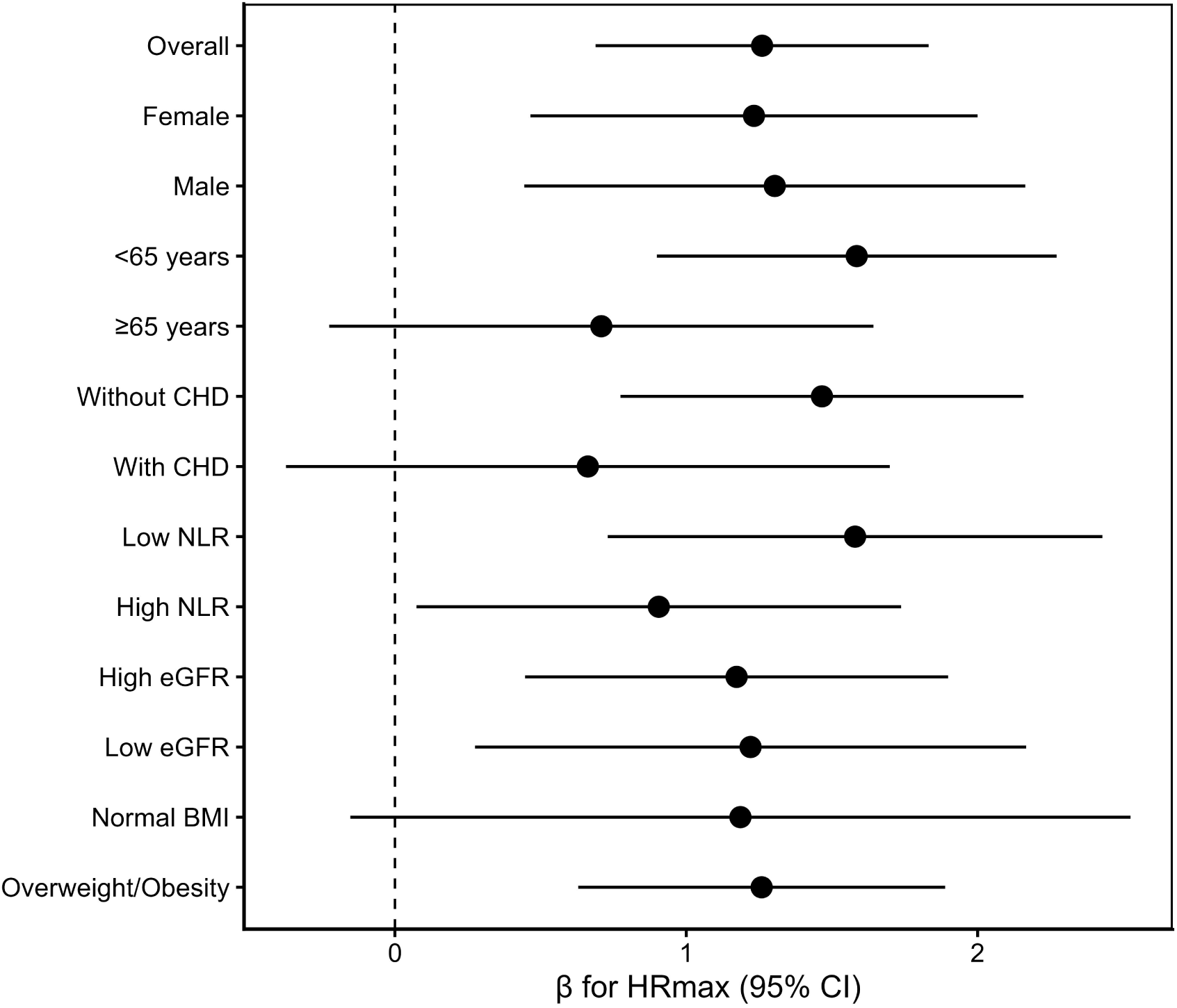
Subgroup analyses of the association between maximum heart rate and 6-minute walk distance. The forest plot shows regression coefficients and corresponding 95% confidence intervals across predefined and exploratory subgroups. No statistically significant interaction was observed for sex, age group, CHD,NLR, eGFR, or BMI. Abbreviations:CHD, coronary heart disease;CHF, congestive heart failure;NLR, neutrophil-to-lymphocyte ratio;eGFR, estimated glomerular filtration rate;BMI, body mass index.

Sensitivity analysis demonstrated good model robustness, and the results of the 10-fold cross-validation were consistent with those of the primary analysis (Supplementary Table S3).

## Discussion

In this cross-sectional study, we systematically evaluated the associations between multiple exercise-derived ICG hemodynamic parameters and exercise capacity in patients with hypertension. After adjustment for demographic characteristics, comorbidities, hemodynamic parameters, and laboratory measurements, HRmax showed a consistent association with 6MWD. This association remained consistent in both the linearity analysis and subgroup analyses.

Notably, among the exercise-derived ICG hemodynamic parameters, only HRmax remained independently associated with 6MWD. This finding suggests that different exercise-derived ICG hemodynamic parameters may provide distinct physiological information regarding exercise capacity in patients with hypertension.

Heart rate is a key physiological determinant of exercise capacity. This relationship can be explained by the Fick principle, whereby the increase in oxygen uptake (VO₂) during exercise depends on an increase in cardiac output, which is determined by both heart rate and stroke volume. Therefore, the ability of heart rate to increase appropriately in response to metabolic demand during exercise—that is, an adequate heart rate response—is critical for maintaining oxygen delivery and exercise performance. Previous studies have consistently shown that chronotropic incompetence (CI) limits the normal increase in heart rate during exercise, thereby reducing cardiac output reserve, peak oxygen uptake (peak VO₂), and exercise capacity^[12,13]^.

Efthimiadis et al. reported that heart rate reserve was closely associated with peak VO₂^[14]^. Chua et al. demonstrated that improvement in chronotropic function was accompanied by increases in both peak heart rate and peak VO₂^[15]^. Kagami et al. further showed that an adequate heart rate response contributes to the maintenance of cardiac output during exercise^[16]^. Although these studies were not all conducted in patients with hypertension, they consistently support the physiological relationship between heart rate response and exercise capacity across different disease populations and exercise assessment settings.

Although both heart rate and stroke volume contribute to the regulation of cardiac output during exercise, our findings suggest that these two components may not provide equivalent information for distinguishing differences in exercise capacity. Different ICG-derived hemodynamic parameters reflect distinct aspects of circulatory reserve and, therefore, may differ in their associations with exercise capacity.

Previous studies have shown that the relative contributions of heart rate and stroke volume to the increase in cardiac output vary according to exercise intensity. Using continuous impedance cardiography during 6MWT, Liu et al. demonstrated that the contribution of heart rate to the increase in cardiac output progressively increased as exercise continued^[8]^. Franzoni et al. further observed that when stroke volume reserve was limited because of impaired cardiac function, a compensatory increase in heart rate became an important mechanism for maintaining oxygen delivery^[17]^. In contrast, stroke volume is influenced by multiple physiological factors and therefore exhibits substantial interindividual variability, with responses ranging from an early plateau to a continued increase or even a decline during exercise^[18]^. Rowland et al. likewise reported considerable heterogeneity in stroke volume responses across different populations^[19]^.

Therefore, the finding that HRmax was the only exercise-derived ICG hemodynamic parameter independently associated with 6MWD in the present study does not imply that heart rate is more important than stroke volume. Rather, it suggests that, during the 6MWT in patients with hypertension, heart rate–related parameters may more consistently reflect differences in circulatory reserve associated with exercise capacity than stroke volume–related parameters.

Although HRmax remained consistently associated with exercise capacity in the present study, the mechanisms underlying this association could not be directly determined. Previous evidence suggests that impaired autonomic regulation may represent one of the mechanisms influencing heart rate response during exercise.

Maciorowska et al. reported that heart rate variability was closely associated with hemodynamic status in patients with hypertension^[20]^. As heart rate variability is an established indicator of autonomic regulation, these findings suggest that altered autonomic regulation may contribute to impaired circulatory function in patients with hypertension. Masroor et al. further demonstrated that exercise training improved heart rate response in parallel with improvements in autonomic function^[21]^. In addition, Prasad et al. suggested that chronotropic dysfunction may precede the clinical onset of hypertension^[22]^.

Taken together, these findings suggest that a limited HRmax observed in the present study may not simply reflect an inadequate increase in heart rate during exercise, but may also indicate impaired chronotropic function potentially related to autonomic dysregulation, thereby limiting the mobilization of circulatory reserve during exercise.

Kurpaska et al. applied exercise impedance cardiography and demonstrated abnormal hemodynamic responses during exercise in patients with hypertension and exertional dyspnea, which may be associated with reduced exercise capacity^[23]^. Their subsequent study further suggested that multiple exercise-derived ICG hemodynamic parameters were associated with exercise capacity^[5]^. Building upon these findings, the present study further compared the relative associations of different ICG-derived parameters and demonstrated that HRmax showed the most consistent association with 6MWD.

On the other hand, although the study populations and exercise endpoints differed from those of the present study, previous investigations by Efthimiadis, Chua, and Kagami have consistently supported the association between heart rate–related parameters and exercise capacity from different perspectives, including heart rate reserve, improvement in chronotropic function, and the contribution of heart rate to cardiac output during exercise^[14–16]^. By using 6MWD as the exercise capacity endpoint, the present study demonstrated similar findings, suggesting that this association may extend across different exercise assessment approaches.

In addition, age and sex have been identified as major determinants of 6MWD in the prediction model developed by Ramos et al., and consistent associations were observed in the present study^[24]^.

Our findings suggest that, when exercise capacity is assessed using ICG combined with the 6MWT, particular attention should be paid to heart rate response during exercise. HRmax may not only reflect the immediate heart rate response to exercise but may also provide additional information regarding overall circulatory reserve and chronotropic function, thereby complementing the assessment of exercise capacity. The European Association of Preventive Cardiology (EAPC) position statement recommends that peak heart rate and heart rate reserve remain important indicators for exercise intensity assessment when CPET is unavailable^[25]^. Although the 6MWT is a submaximal exercise test and HRmax cannot replace CPET-derived parameters, the combination of ICG and the 6MWT may serve as a practical complementary approach for the assessment of exercise capacity in settings where CPET is not readily available.

This study has several limitations. First, this was a single-center retrospective cross-sectional study, which precludes causal inference. Second, the sample size was relatively small, and the findings require validation in larger, multicenter populations. Third, CPET was not performed; therefore, peak VO₂ and other metabolic parameters were unavailable. Fourth, autonomic function was not directly assessed, and the proposed mechanistic interpretation remains indirect. Finally, no follow-up data were available, precluding evaluation of the prognostic significance of the observed associations. Therefore, the findings of the present study should be interpreted with caution, and their clinical applicability warrants further investigation in prospective multicenter studies.

## Conclusion

Among multiple exercise-derived ICG hemodynamic parameters in patients with hypertension, HRmax remained independently associated with 6MWD and demonstrated the most consistent association. These findings suggest that HRmax may represent an important dynamic hemodynamic parameter that warrants particular attention when exercise capacity is assessed using ICG combined with the 6MWT. The combination of ICG and 6MWT may provide a simple and accessible complementary approach for evaluating exercise capacity.

## Data Availability

The datasets generated and analyzed during the current study are available from the corresponding author upon reasonable request.

## Ethics Approval

This study was approved by the Ethics Committee of The Second Affiliated Hospital of Nanjing Medical University(Approval No. 2026-KY-285-01). The study was conducted in accordance with the principles of the Declaration of Helsinki.

## Author Contributions

Ting Yu designed the study, collected the data, performed the statistical analyses, and drafted the manuscript.

Guozhong Ji supervised the study and critically revised the manuscript. All authors reviewed and approved the final manuscript.

